# A staged screening protocol identifies people with biomarkers related to neuronal alpha-synuclein disease

**DOI:** 10.1101/2024.06.03.24308229

**Authors:** Ethan G. Brown, Lana M. Chahine, Andrew Siderowf, Caroline Gochanour, Ryan Kurth, Micah J. Marshall, Chelsea Caspell-Garcia, Michael C. Brumm, Christopher S. Coffey, Craig E. Stanley, Monica Korell, Bridget McMahon, Maggie Kuhl, Kimberly Fabrizio, Laura Heathers, Tatiana M. Foroud, Luis Concha-Marambio, Claudio Soto, Sohini Chowdhury, Tanya Simuni, Kenneth Marek, Caroline M. Tanner, the Parkinson Progression Marker Initiative

## Abstract

**Importance:** Identifying individuals in the earliest stages of synucleinopathy is essential to evaluate drugs aimed to slow progression or prevent manifest disease. Remote identification of hyposmic individuals may enable scalable recruitment of participants with underlying alpha-synuclein pathology.

**Objective:** To evaluate the performance of a staged screening paradigm using smell testing to enrich for deficit on dopaminergic transporter (DAT) imaging and pathologic alpha-synuclein aggregation.

**Design:** Cross-sectional analysis of data from the Parkinson’s Progression Markers Initiative (PPMI).

**Setting:** Screening activities were completed both at home and local PPMI sites.

**Participants:** Individuals aged 60 and older without a diagnosis of Parkinson’s disease **Interventions or Exposures:** Participants were asked to complete a University of Pennsylvania Smell Identification Test (UPSIT) remotely. Participants with hyposmia were invited to complete DAT imaging, which determined eligibility for enrollment in longitudinal assessments and further biomarker evaluation including cerebrospinal fluid synuclein seed amplification assay (synSAA).

**Main Outcomes and Measures:** We determined the proportion of people with hyposmia, impaired DAT binding, and positive synSAA and explored determinants of these biomarkers.

**Results:** As of January 29, 2024, 49,843 participants were sent an UPSIT and 31,293 (63%) completed it. Of UPSIT completers, 8,301 (27%) scored <15^th^ %ile. Of 1,546 who completed DAT, 19% had DAT binding < 65% expected for age and sex. Self-reported features were independently associated with severe hyposmia (UPSIT <10^th^ %ile), such as REM sleep behavior disorder (RBD) or dream enactment behavior (DEB) (aOR: 1.9, 95% CI 1.7–2.1) and subjective smell loss (aOR: 15.0, 95% CI 13.7–16.3). Participants with an UPSIT <10^th^ %ile (N=1,221) had greater likelihood of low DAT binding compared to participants with an UPSIT in the 10^th^ – 15^th^ %ile (OR 3.01, 95% CI 1.85–4.91). Among remotely recruited participants with synSAA results obtained, 198/363 (55%) had positive synSAA at baseline. This proportion increased when the cohort was limited to an UPSIT<10^th^ %ile (182/257, 71%).

**Conclusion and Relevance:** Remote screening for severe hyposmia identifies participants with a high proportion of positive synSAA and reduced DAT binding. This staged screening protocol is an effective approach to identify cohorts for therapeutic trials aiming to slow progression in alpha-synucleinopathy.

## Introduction

Early recognition of neurodegenerative conditions related to alpha-synuclein (aSyn) aggregation, or synucleinopathies, may be an essential step toward identifying effective therapies. Recent discoveries have enabled identification of people with misfolded aSyn, even prior to the onset of symptoms or disability.^1-3^ However, as these conditions are relatively uncommon, large-scale screening efforts are necessary to identify people with biomarkers that suggest the presence of these conditions.

The Parkinson’s Progression Markers Initiative (PPMI)^4^ has implemented a staged screening protocol, beginning with non-invasive, easily administered, and scalable assessments then gradually increasing specificity to identify people with biomarkers consistent with synuclein-related neurologic disease. Recent biomarker data from the PPMI study has enabled a biologic definition of Neuronal Synuclein Disease (NSD), a new terminology encompassing Parkinson’s disease (PD) and Dementia with Lewy Bodies (DLB), to describe individuals with biomarker evidence of neuronal synucleinopathy. Findings have also supported an integrated biomarker and clinical staging platform called the NSD-ISS.^5^ Stages 1 and 2 in the NSD-ISS are individuals with biomarkers of synuclein and dopamine dysfunction without meaningful functional impairment.

Our staged screening protocol is designed to identify individuals in NSD Stage 2 (detectable biomarkers with clinical features but no functional impairment) using hyposmia to enrich for presence of pathologic synuclein aggregation (measured by the aSyn seed amplification assay [SAA]) and dopamine dysfunction (measured by dopamine transporter imaging [DAT]). We describe the performance characteristics of this staged screening protocol and highlight features that may further improve its efficiency to identify an early-stage NSD study cohort.

## Methods

### Overview of study

PPMI is a longitudinal study sponsored by The Michael J. Fox Foundation for Parkinson’s Research (MJFF) that seeks to improve the treatment of PD and related conditions by identifying biomarkers of disease onset and progression.^6,7^ PPMI enrolled varied cohorts from 2010 – 2018. Since 2020, PPMI has had a major focus on enrollment and longitudinal follow-up of participants with features indicative of synucleinopathy but without a clinical diagnosis of PD or DLB. This paper focuses on the subset of these participants that are recruited remotely and go through a staged screening process to determine eligibility.

### Recruitment for staged screening

Participants were recruited for staged screening through three different remote pathways. From October 2020 through October 2021, participants were identified through an online questionnaire hosted by the MJFF website. In July 2021, an online longitudinal study involving participant reported outcomes (PROs), open to anyone age 18 years or older with or without PD, was launched as PPMI Online.^8,9^ Starting November 2021, participants already enrolled in PPMI Online who met criteria (see below) were invited to enter staged screening. Starting May 10, 2022, recruitment strategies were broadened to directly invite community members to complete a smell test through a simplified recruitment pathway called Smell Test Direct (ST Direct).

Within these centralized pathways, campaigns were flexible and could target aged individuals with features related to synucleinopathy, such as a diagnosis of RBD, dream enactment behavior, and loss of sense of smell. Potential participants identified locally by PPMI site investigators and coordinators undergo a separate screening procedure, not described in this analysis.^8^ While the MJFF screener is no longer active, PPMI Online and ST Direct remain active methods of recruitment pathways to identify participants for PPMI enrollment. At the time of writing, PPMI Online is only available in the United States, while ST Direct is active in the United States, Canada, the United Kingdom, and Europe.

### Online questionnaire

Participants recruited through PPMI and ST Direct completed questions about features related to synucleinopathy before completing a smell test. Questions include (1) whether a healthcare professional had ever diagnosed RBD, (2) whether the participant had a suspicion or had been told that they acted out their dreams,^10^ (3) whether or not they had noticed problems with their sense of smell, and (4) whether they had a first degree relative who was diagnosed with Parkinson’s disease. Participants of PPMI Online completed other participant-reported outcomes (PROs) related to motor and non-motor symptoms. All questions were recorded but did not play a role in determining eligibility for further screening.

### Staged screening methodology

Participants identified through any recruitment pathway who (i) did not have a diagnosis of PD and (ii) were 60 years old or older were mailed a University of Pennsylvania Smell Identification Test (UPSIT) that they could complete at home, entering answers online. Initially participants were considered hyposmic if scores were less than the 15^th^ %ile compared to normative values for age and sex.^11^ To more efficiently identify individuals with active synucleinopathy seeds and dopamine dysfunction, coupled with preliminary analysis of the screening performance, the threshold for hyposmia was decreased to the 10^th^ %ile starting January 6, 2023.

Participants identified as hyposmic were contacted for additional phone screening by a centralized team to confirm eligibility to undergo DAT. This process confirmed there were no disqualifying medical conditions or medications – including antipsychotic medications or medications that would interfere with DAT. Eligible and agreeable participants were invited to complete a DAT at a PPMI site.^12^ The lowest putamen specific binding ratio (SBR) was determined and compared to age- and sex-adjusted expected values. Participants with a lowest putamen SBR <80% expected for age and sex were considered eligible and invited to full clinical enrollment. Preliminary analysis of the staged screening performance showed that participants with lower age and sex expected putamen SBR were more likely to have symptoms and signs related to synucleinopathy (see *Results*, Table 3). To capture more participants in an even earlier stage of disease, we increased the threshold for eligibility to ≤100% age and sex expected putamen SBR on May 10 2022.

### PPMI in-person evaluations

Once enrolled at a site, PPMI participants underwent in-person visits.^8^ Clinical evaluations involved standardized PROs and assessments covering both motor and non-motor symptoms, as well as neuropsychological testing.^6^ Clinical assessments used for this analysis were taken from the baseline visit and include Parts I – III of the Movement Disorders Society Unified Parkinson’s Disease Rating Scale (MDS-UPDRS),^13^ the Montreal Cognitive Assessment (MoCA),^14^ the Scales for Outcomes in Parkinson’s Disease – Autonomic (SCOPA-Aut),^15^ and the REM Sleep Behavior Disorder Screening Questionnaire (RBDSQ).^16^ Biological assessments include blood and urine collection, skin biopsy, and cerebrospinal fluid (CSF) analysis. CSF is sent for alpha-synuclein seed amplification assay (synSAA) through Amprion Inc.^1,2,17^ Potential results from the Amprion assay include Type 1, most commonly seen in Lewy Body disease, Type 2, most commonly seen in multiple system atrophy (MSA), negative, or inconclusive. Imaging assessments include DAT and MRI imaging.

### Statistical Analysis

Data was downloaded for analysis on January 29 2024. Only participants that completed the UPSIT remotely were included. We first determined the number of participants included and excluded at each stage of the screening process and used descriptive statistics to compare groups. We compared participants who completed each step with those who were eligible but did not complete each step along the screening pathway. Univariate statistics were estimated using Chi-square and Fisher’s Exact tests for categorical variables and t-tests for continuous variables. We used odds ratio to determine the relationship between severe and moderate hyposmia (UPSIT <10^th^ compared to 10^th^–15^th^ percentile for age and sex) and severe and moderate reduction in DAT binding (lowest putamen SBR <65% compared to 65-100% expected for age and sex). We chose 65% to represent substantially reduced DAT binding based on prior literature showing a high association with a clinical diagnosis of PD. We implemented a linear regression analysis adjusting for age and sex to determine the relationship between DAT SBR <65% and MDS-UPDRS Part III among hyposmic individuals with DAT results. Among participants who had completed baseline assessments, we compared summary scores of these measures between those with an SBR of <65% compared to an SBR of 65-100% and people with synSAA Type1 seeds compared to negative synSAA. Univariate and multivariate regression models were used to determine the association of features collected through self-report and UPSIT. We adjusted for age of UPSIT completion and sex, since both these variables can influence smell^18^ and DAT binding.^19^ Finally, we used odds ratio to determine the association between UPSIT (<10^th^ %ile vs 10^th^–15^th^ %ile) and synSAA Type 1. All analyses were performed using SAS v9.4 (SAS Institute Inc., Cary, NC; sas.com; RRID:SCR 008567).

## Results

### Smell testing

As of January 29 2024, 49,843 participants were invited to complete remote smell tests and 31,293 (63%) completed them (Figure 1; Table 1). The largest volume of participants to complete the smell test came through ST Direct (n = 24,921), though the highest rate of UPSIT completion was from PPMI Online (84%; eFigures 1-3). Demographics were similar between people who completed a smell test and people who were provided but never completed a test (eTable 1). Among participants who did return the UPSIT, the average result was the 29^th^ %ile with 8,301 (27%) scoring at or below the 15^th^ %ile and 4,639 (15%) below the 10^th^ %ile (Table 1).

**Figure 1.**
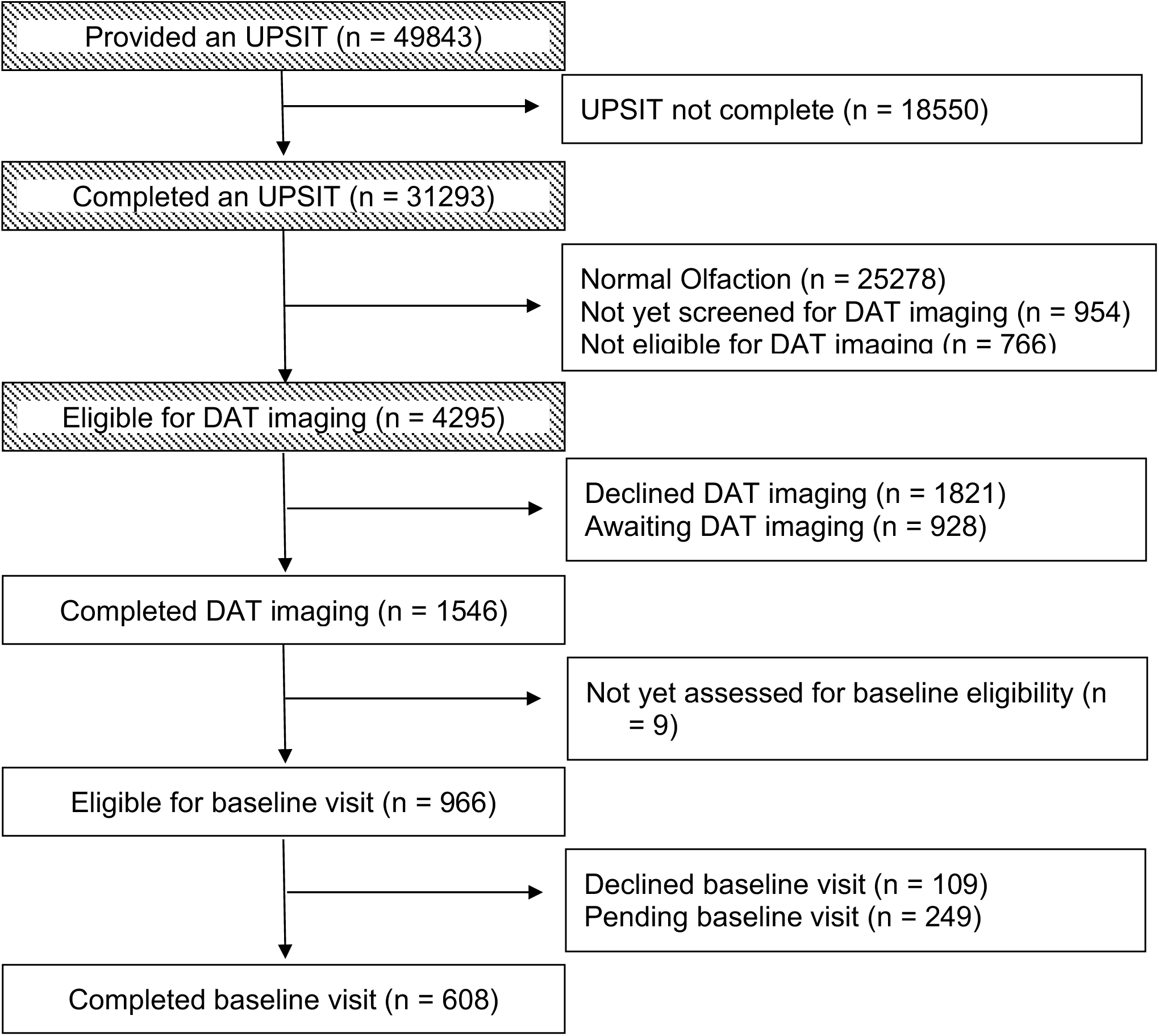
Flowchart of participants remotely screened for the PPMI Prodromal Cohort as of Jan 29, 2024. Cross hatch squares indicate activities that are completed remotely.

**Table 1.** Characteristics of participants at various stages of the screening protocol for the Prodromal Cohort of PPMI.

|  | Participants with a completed UPSIT |  |  |  |
| --- | --- | --- | --- | --- |
|  | Participants with a completed UPSIT<br>(N = 31293) | Participants with a completed DaTScan<br>(N = 1546) | Participants with a completed DaTScan |  |
|  |  |  | Participants with SBR < 65%<br>(N = 294) | Participants with SBR ≥ 65%<br>(N = 1252) |
| **Age at UPSIT (years), Mean (SD)** | 68.3 (6.0) | 67.7 (5.5) | 68.4 (5.3) | 67.5 (5.5)<sup>^</sup> |
| Median (Min, Max) | 67.4 (23.6, 95.9) | 66.9 (59.3, 87.2) | 68.1 (59.6, 87.0) | 66.6 (59.3, 87.2) |
| **Sex, n (%)** |  |  |  |  |
| Female | 22131 (71%) | 1001 (65%) | 165 (56%)** | 836 (67%)<sup>†</sup> |
| Male | 9162 (29%) | 545 (35%) | 129 (44%) | 416 (33%) |
| **UPSIT Percentile, Mean (SD)** | 29.3 (17.8) | 6.6 (4.0) | 5.2 (3.2)** | 6.9 (4.2)<sup>†</sup> |
| Median (Min, Max) | 27.0 (1.0, 93.5) | 6.0 (1.0, 15.0) | 5.0 (1.0, 15.0) | 6.0 (1.0, 15.0) |
| **UPSIT Percentile Categories, n (%)** |  |  |  |  |
| < 10th Percentile | 4639 (15%) | 1221 (79%) | 271 (92%) | 950 (76%)<sup>^</sup> |
| ≥ 10th and ≤ 15th Percentile | 3662 (12%) | 325 (21%) | 23 (8%) | 302 (24%) |
| > 15th Percentile | 22992 (73%) | 0 | 0 | 0 |
| **SBR % Expected, Mean (SD)** | - | 89.5 (30.8) | 48.9 (12.2) | 99.1 (25.7) |
| Median (Min, Max) | - | 86.7 (14.3, 209.1) | 52.0 (14.3, 64.9) | 93.1 (65.0, 209.1) |
| **SBR Categories, n (%)** |  |  |  |  |
| <65% | - | 294 (19%) | 294 (100%) | 0 |
| 65% - < 80% | - | 306 (20%) | 0 | 306 (24%) |
| 80% - ≤ 100% | - | 460 (30%) | 0 | 460 (37%) |
| 100% + | - | 486 (31%) | 0 | 486 (39%) |
| **MDS-UPDRS Total\*, Mean (SD)** | - | 13.2 (10.4) | 17.0 (12.3) | 11.9 (9.3)<sup>^</sup> |
| Median (Min, Max) | - | 11.0 (0.0, 69.0) | 14.0 (0.0, 69.0) | 9.0 (0.0, 63.0) |
| Not evaluated | - | 941 | 132 | 809 |
| **MDS-UPDRS Part I, Mean (SD)** | - | 6.8 (5.0) | 7.9 (5.6) | 6.4 (4.8)<sup>^</sup> |
| Median (Min, Max) | - | 6.0 (0.0, 31.0) | 7.0 (0.0, 26.0) | 5.0 (0.0, 31.0) |
| Not evaluated | - | 918 | 125 | 793 |
| **MDS-UPDRS Part II, Mean (SD)** | - | 2.1 (3.5) | 3.2 (4.3) | 1.8 (3.1)<sup>^</sup> |
| Median (Min, Max) | - | 1.0 (0.0, 25.0) | 2.0 (0.0, 24.0) | 0.0 (0.0, 25.0) |
| Not evaluated | - | 911 | 123 | 788 |
| **MDS-UPDRS Part III, Mean (SD)** | - | 4.5 (5.3) | 6.3 (6.6) | 3.8 (4.6)<sup>^</sup> |
| Median (Min, Max) | - | 3.0 (0.0, 35.0) | 4.0 (0.0, 35.0) | 2.0 (0.0, 30.0) |
| Not evaluated | - | 923 | 124 | 799 |

\*MDS-UPDRS scores are from the baseline clinical visit.
Assessments between participants with lowest putamen SBR < 65% expected for age and sex and lowest putamen SBR ≥ 65% expected for age and sex were compared using Chi-square and Fisher's Exact tests for categorical variables and t-tests for continuous variables. <sup>^</sup> indicates p-value < 0.01; <sup>†</sup> indicates p-value < 0.001.

### DAT imaging

A total of 4,295 participants were screened and deemed eligible for DAT imaging; 2,474 (58%) agreed to participate and 1,546 (36%) completed DAT imaging thus far (Table 1). Participants who declined DAT imaging were more likely to be female and older compared to completers (eTable 2). Among those who completed DAT imaging, 294 (19%) had a lowest putamen SBR below 65% age and sex expected, whereas 306 (20%) had a lowest putamen SBR between 65% and 80% expected (Table 1). Among hyposmic individuals who were eligible for DAT imaging to be completed, and after adjusting for age and sex, people with a lowest putamen SBR <65% had a 2.05 higher MDS-UPDRS Part III score (95% CI 1.14 – 2.97) compared to people with a lowest putamen SBR 65-100%; MDS-UPDRS total and other sub-scores were also significantly different (Table 1).

### Relationship between DAT imaging and hyposmia

To determine how hyposmia was related to DAT, we restricted our analysis to a subsample of people who were eligible based on an UPSIT threshold of ≤15^th^ %ile (i.e. participants who completed the UPSIT before January 6^th^, 2023). Among those 742 participants, 420 (57%) had an UPSIT <10^th^ %ile and 322 (43%) had an UPSIT in the 10^th^ – 15^th^ %ile. People with UPSIT <10^th^ %ile had three times the odds of having a lowest putamen SBR <65 % expected compared to people with UPSIT in the 10^th^ – 15^th^ %ile (79/420 [19%] vs 23/322 [7%], OR: 3.01, 95% CI 1.85 – 4.91). The distribution of UPSIT and DAT result is shown in eFigure 4.

### Alpha-synuclein seed amplification assay

Among the 1,546 participants who received a DaTScan, 857 were eligible for and 608 have enrolled in PPMI Clinic. CSF synSAA results in 363 of these participants were available by the time of this analysis: 198 (55%) were positive with a Type 1 pattern, 1 (0.3%) was positive with a Type 2 pattern, 4 (1.1%) were inconclusive and 160 (44%) were negative. Among participants who had an UPSIT <10^th^ %ile with synSAA, 182/257 (71%) had an Type 1 pattern (Figure 2).

**Figure 2:**
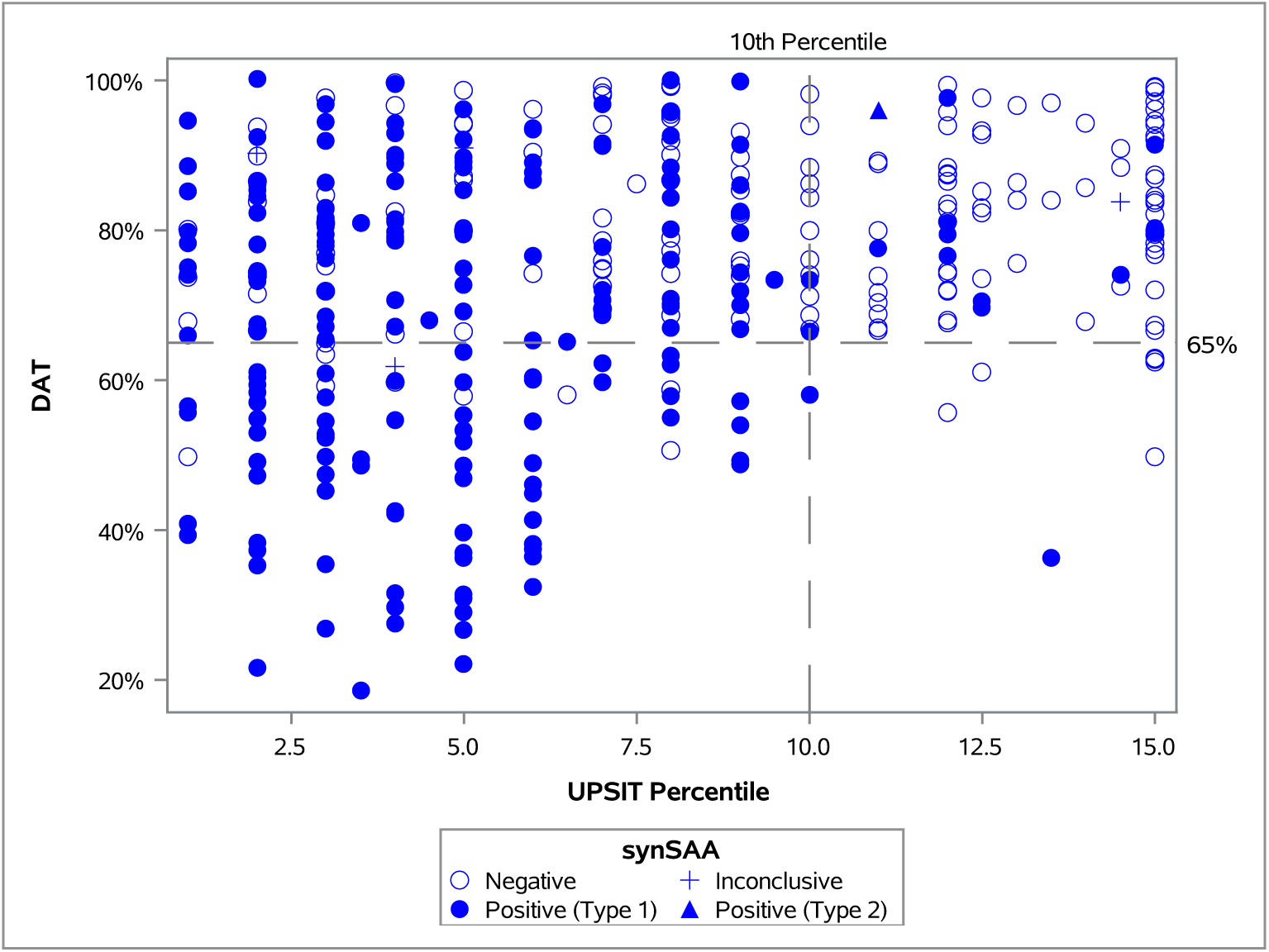
Relationship between UPSIT results, DAT imaging results, and CSF synSAA results among participants recruited through remote staged screening for PPMI and enrolled in the Prodromal Cohort of PPMI.

### Determinants of hyposmia

We determined the association between the responses to questions in PPMI Online and ST Direct (eTable 3) and severe hyposmia, which we defined as UPSIT <10^th^ %ile given the association with DaTScan and synSAA results. Having an RBD diagnosis, reporting dream enactment behavior (DEB) even without an RBD diagnosis, or reporting loss of sense of smell were associated with an UPSIT <10^th^ %ile (Table 3). In multivariate analysis, significant predictors of hyposmia included having a diagnosis of RBD or reporting DEB (aOR: 1.9, 95% CI 1.7 – 2.2, p<0.001) and a self-report of hyposmia (aOR: 15.0, 95% CI 13.7 – 16.3, p<0.001).

### Clinical and biological correlates of abnormal DAT imaging and aSyn-SAA

Among people with synSAA results, participants with Type 1 synSAA were older, more likely to be male and have lower UPSIT. People with severe hyposmia (UPSIT <10^th^ %ile) were more likely to have Type 1 synSAA compared to moderate hyposmia (UPSIT 10^th^ – 15^th^ %ile; OR: 12.9, 95% CI 7.1 – 23.4; Figure 2). The total score on the RBD Screening Questionnaire was higher in people with Type 1 synSAA compared to people with negative synSAA. Participants with Type 1 CSF aSyn have a lower percent of age and sex expected putamen SBR than people with negative synSAA (Table 2). Out of 91 participants with lowest putamen SBR <65%, 77 (85%) had Type 1 synSAA (Table 2). Other clinical assessments were not significantly different after adjustment for age and sex (Table 2).

**Table 2.** CSF aSyn SAA results among participants who completed an UPSIT remotely and were ultimately eligible a baseline visit in the Prodromal Cohort of PPMI.

|  | aSyn SAA Type 1,<br>(N=198) | aSyn SAA Negative,<br>(N=160) | P-value** | P-value(adjusted)*** |
| --- | --- | --- | --- | --- |
| **Age at UPSIT (years), Mean (SD)** | 68.1 (5.0) | 65.4 (4.7) | <.0001 |  |
| Median (Min, Max) | 67.7 (60.0, 84.2) | 64.3 (59.5, 85.9) |  |  |
| **Sex, n (%)** |  |  | <.0001 |  |
| Female | 99 (50%) | 128 (80%) |  |  |
| Male | 99 (50%) | 32 (20%) |  |  |
| **UPSIT Percentile, Mean (SD)** | 5.3 (3.1) | 9.5 (4.2) | <.0001 |  |
| Median (Min, Max) | 5.0 (1.0, 15.0) | 10.0 (1.0, 15.0) |  |  |
| **UPSIT Percentile Categories, n (%)** |  |  | <.0001 |  |
| ≥ 10<sup>th</sup> and ≤ 15th Percentile | 16 (8%) | 85 (53%) |  |  |
| < 10% | 182 (92%) | 75 (47%) |  |  |
| **SBR % Expected, Mean (SD)** | 67.7 (19.7) | 80.9 (12.1) | <.0001 |  |
| Median (Min, Max) | 70.3 (18.5, 100.2) | 82.0 (49.7, 99.7) |  |  |
| **SBR Categories, n (%)** |  |  | <.0001 |  |
| < 65% | 77 (39%) | 14 (9%) |  |  |
| 65% - < 80% | 60 (30%) | 59 (37%) |  |  |
| 80% - ≤ 100% | 61 (31%) | 87 (54%) |  |  |
| **MDS-UPDRS Total\*, Mean (SD)** | 14.2 (11.8) | 11.4 (9.2) | 0.0166 | 0.0881 |
| Median (Min, Max) | 11.0 (0.0, 69.0) | 9.0 (0.0, 63.0) |  |  |
| Not evaluated | 13 | 8 |  |  |
| **MDS-UPDRS Part I, Mean (SD)** | 6.9 (5.1) | 6.2 (4.9) | 0.1860 | 0.1277 |
| Median (Min, Max) | 5.0 (0.0, 26.0) | 5.0 (0.0, 31.0) |  |  |
| Not evaluated | 6 | 3 |  |  |
| **MDS-UPDRS Part II, Mean (SD)** | 2.4 (4.0) | 1.7 (3.2) | 0.0552 | 0.2703 |
| Median (Min, Max) | 1.0 (0.0, 25.0) | 0.0 (0.0, 20.0) |  |  |
| Not evaluated | 5 | 0 |  |  |
| **MDS-UPDRS Part III, Mean (SD)** | 5.1 (6.1) | 3.6 (4.2) | 0.0092 | 0.2152 |
| Median (Min, Max) | 3.0 (0.0, 35.0) | 2.0 (0.0, 21.0) |  |  |
| Not evaluated | 7 | 5 |  |  |
| **RBDSQ Total, Mean (SD)** | 5.2 (4.2) | 3.0 (2.7) | <.0001 | <.0001 |
| Median (Min, Max) | 4.0 (0.0, 13.0) | 2.0 (0.0, 12.0) |  |  |
| Not evaluated | 3 | 0 |  |  |
| **SCOPA-AUT Total, Mean (SD)** | 10.6 (6.4) | 9.9 (7.1) | 0.2935 | 0.7141 |
| Median (Min, Max) | 9.0 (0.0, 36.0) | 8.0 (0.0, 41.0) |  |  |
| Not evaluated | 3 | 0 |  |  |
| **MoCA Total, Mean (SD)** | 26.8 (2.2) | 27.1 (2.2) | 0.3008 | 0.5899 |
| Median (Min, Max) | 27.0 (21.0, 30.0) | 28.0 (21.0, 30.0) |  |  |
| Not evaluated | 9 | 5 |  |  |

\*MDS-UPDRS scores are from the baseline clinical visit.
\*\*P-values were obtained using Chi-square and Fisher's Exact tests for categorical variables and t-tests for continuous variables.
\*\*\* Age and Sex adjusted P-values were obtained using linear regression.

**Table 3.** Relationship between responses to screening questionnaire and severe hyposmia among participants screened via ST-Direct or PPMI Online.

| Characteristic | UPSIT ≥ 10th %ile | UPSIT < 10th %ile | OR | 95% CI | Total | N. missing, PNTA or Not Sure |
| --- | --- | --- | --- | --- | --- | --- |
| **RBD Diagnosis, n(%)** | 555 (1.97%) | 400 (1.42%) | 4.53 | (3.97, 5.18) | 28144 | 1616 |
| **DEB without RBD\*, n(%)** | 2598 (9.22%) | 936 (3.32%) | 2.34 | (2.15, 2.54) | 28173 | 1587 |
| **Self-reported hyposmia, n(%)** | 2380 (9.5%) | 2503 (9.99%) | 16.28 | (15.02, 17.65) | 25059 | 4701 |
| **Family History of PD, n(%)** | 6090 (21.66%) | 933 (3.32%) | 0.86 | (0.79, 0.93) | 28113 | 1647 |

Percents represent proportion of total across row. PNTA: Prefer not to answer.
\*Indicates participants who have not received a diagnosis of RBD but experience dream enactment behavior.

## Discussion

We have demonstrated that a large-scale community staged screening protocol based on age and hyposmia can successfully identify a broad cohort with dopaminergic deficit and a positive misfolded aSyn biomarker (CSF synSAA). We expect that hyposmic individuals with positive synSAA are at high risk for progression of striatal neurodegeneration and clinical symptoms.

Longitudinal studies are required to confirm this time course and identify other factors relevant to disease progression. A key finding was that 19% of people with hyposmia had a reduction in DAT (lowest putamen SBR <65% of age- and sex-expected) and 55% of people with hyposmia and DAT binding ≤100% expected were synSAA positive. This represents substantial enrichment, considering the estimated prevalence of positive synSAA in this age group is between 4-8%.^2,20^ Therefore, hyposmia is a potential tool to identify and enroll synSAA positive individuals for clinical trials seeking to test therapies in people with NSD prior to the onset of functionally impactful symptoms.

We have presented the characteristics of participants at each stage of screening from olfactory testing to DAT imaging and CSF collection. Our data support the feasibility of identifying high numbers of individuals with hyposmia, DAT deficit and positive synSAA. Our data also highlight that to be successful, such initiatives will require high volume screening due to attrition at each step of screening, i.e. (i) agreement to DAT imaging at a clinical site (58% of eligible agreed) and (ii) agreement to follow up (89% of eligible consented). The staged screening paradigm, applied at scale, delivered a high volume of individuals enriched with hyposmia, low DAT binding, and synSAA positive status. These data strongly support that this staged risk paradigm can successfully enroll a synSAA positive/DAT deficit biomarker cohort consistent with Stage 2 NSD.^5^

A key feature of this study was to adapt the staged screening paradigm to improve efficiency of identification of biomarker-defined individuals as data was acquired, thereby reducing cost and participant burden. The UPSIT cutoff was changed from 15^th^ %ile to 10^th^ %ile to enhance the likelihood that individuals had DAT deficit and eliminate unnecessary imaging. Similarly, the DAT cutoff was increased to acquire aSyn SAA data across a greater range of DAT results to better inform the temporal sequence of biomarkers. The screening paradigm is designed for this iterative approach and may be further modified in response to additional data. The goal is both to improve efficiency and reduce costs of identifying individuals and to broaden the key biomarker acquisition to enable the earliest detection of synuclein pathology, closer to the start of neurodegeneration.

We present data showing that more than half of hyposmic participants with positive synSAA have DAT between 65% and 100% age- and sex-expected lowest putamen SBR. This finding implies that misfolded aSyn is present prior to substantial striatal degeneration, suggesting a temporal biomarker pattern prior to symptom onset. Our work expands on data from the Parkinson Associated Risk Syndrome Study (PARS) showing that 11% of hyposmic (<15^th^ %ile) participants had a DAT ≤65% age- and sex-expected putamen SBR compared to 0 normosmic DAT deficit participants.^21^ Importantly in PARS the majority of hyposmic synSAA-positive participants had DAT>80% expected putamen SBR at baseline and for up to six years. These data also support a temporal biomarker pattern of synuclein aggregation followed by dopamine dysfunction. We plan to further investigate the temporal pattern of synSAA and DAT with longitudinal data in PPMI. Further, we plan to modify our staged risk paradigm so hyposmic individuals will be tested first with synSAA and then DAT to determine the likelihood of positive synSAA in hyposmic individuals imaging and DAT deficit in hyposmic synSAA-positive individuals.

### Limitations

We recognize that there are several limitations to this study. Our staged screening protocol successfully identified many people with positive synSAA and DAT deficit, but the efficiency of the staged paradigm can be improved. We mailed several thousand UPSITs; while UPSITs are completed remotely, which allowed for centralized sending and scoring and efficient completion, this process is still moderately resource intensive. Future studies may utilize shorter targeted assessment of smell identification. Our staged paradigm currently depends on expensive and invasive biomarker assessments. While CSF synSAA is the current gold standard it is likely that less invasive samples including blood ^22,23^ or skin ^24^ may be used in the future in place of CSF. Including DAT imaging in our screening may have selected for participants more likely to develop motor symptoms; while it is likely that participants with hyposmia and early cognitive impairment would also demonstrate high likelihood of synSAA positivity, these cohorts need to be evaluated in subsequent studies. Similarly, many of the participants identified in this study may have co-pathology with amyloid or p-tau and we have not assessed these biomarkers yet to understand their role in individuals with synucleinopathy. Finally, as screening and enrollment in longitudinal follow-up is ongoing, participants are still progressing through the screening pipeline. Deducing from the current performance of the staged screening process, the number of participants with positive synSAA and DAT deficit from hyposmics identified in our described time period will continue to increase.

In conclusion, our staged screening protocol was successful in identifying a large NSD biomarker defined cohort. These strategies will inform study design for desperately needed clinical trials that will test therapeutics to slow progression and prevent disability in neuronal alpha-synuclein-related neurodegenerative disease. Given the number of participants we identified with DAT binding >65% expected, these data further support the model that hyposmic individuals with positive synSAA are at high risk for progression of striatal neurodegeneration and eventual clinical symptoms. Longitudinal studies are required to confirm the biomarker time course and identify other clinical and biological factors relevant to disease progression.

## Data Sharing

Data used in the preparation of this article were obtained on January 29, 2024 from the Parkinson’s Progression Markers Initiative (PPMI) database (www.ppmi-info.org/access-data-specimens/download-data), RRID:SCR 006431. For up-to-date information on the study, visit www.ppmi-info.org. This analysis was conducted by the PPMI Statistics Core and used actual dates of activity for participants, a restricted data element not available to public users of PPMI data. Statistical analysis codes used to perform the analyses in this article are shared on Zenodo (10.5281/zenodo.11391274). This analysis used DaTscan and αSyn-SAA results for prodromal participants, obtained from PPMI upon request after approval by the PPMI Data Access Committee.

## Acknowledgment

PPMI – a public-private partnership – is funded by the Michael J. Fox Foundation for Parkinson’s Research and funding partners, including 4D Pharma, Abbvie, AcureX, Allergan, Amathus Therapeutics, Aligning Science Across Parkinson’s, AskBio, Avid Radiopharmaceuticals, BIAL, BioArctic, Biogen, Biohaven, BioLegend, BlueRock Therapeutics, Bristol-Myers Squibb, Calico Labs, Capsida Biotherapeutics, Celgene, Cerevel Therapeutics, Coave Therapeutics, DaCapo Brainscience, Denali, Edmond J. Safra Foundation, Eli Lilly, Gain Therapeutics, GE HealthCare, Genentech, GSK, Golub Capital, Handl Therapeutics, Insitro, Jazz Pharmaceuticals, Johnson & Johnson Innovative Medicine, Lundbeck, Merck, Meso Scale Discovery, Mission Therapeutics, Neurocrine Biosciences, Neuron23, Neuropore, Pfizer, Piramal, Prevail Therapeutics, Roche, Sanofi, Servier, Sun Pharma Advanced Research Company, Takeda, Teva, UCB, Vanqua Bio, Verily, Voyager Therapeutics, the Weston Family Foundation and Yumanity Therapeutics. *Please see the online supplement for a full list of PPMI Authors.

## Author Contributions

*Concept and design:* Brown, Chahine, Siderowf, Foroud, Simmuni, Marek, Tanner

*Acquisition, analysis, or interpretation of data:* All authors.

*Drafting of the manuscript:* Brown, Chahine.

*Critical revision of the manuscript for important intellectual content:* All authors.

*Statistical analysis:* Gochanour, Kurth, Marshall, Caspell-Garcia, Brumm, Coffey.

*Administrative, technical, or material support:* Stanley, Korell, McMahon, Kuhl, Fabrizio, Heathers, Concha-Marambio.

*Supervision:* Siderowf, Coffey, Foroud, Soto, Chowdhury, Simuni, Marek, Tanner.

---

**eFigure 1.**
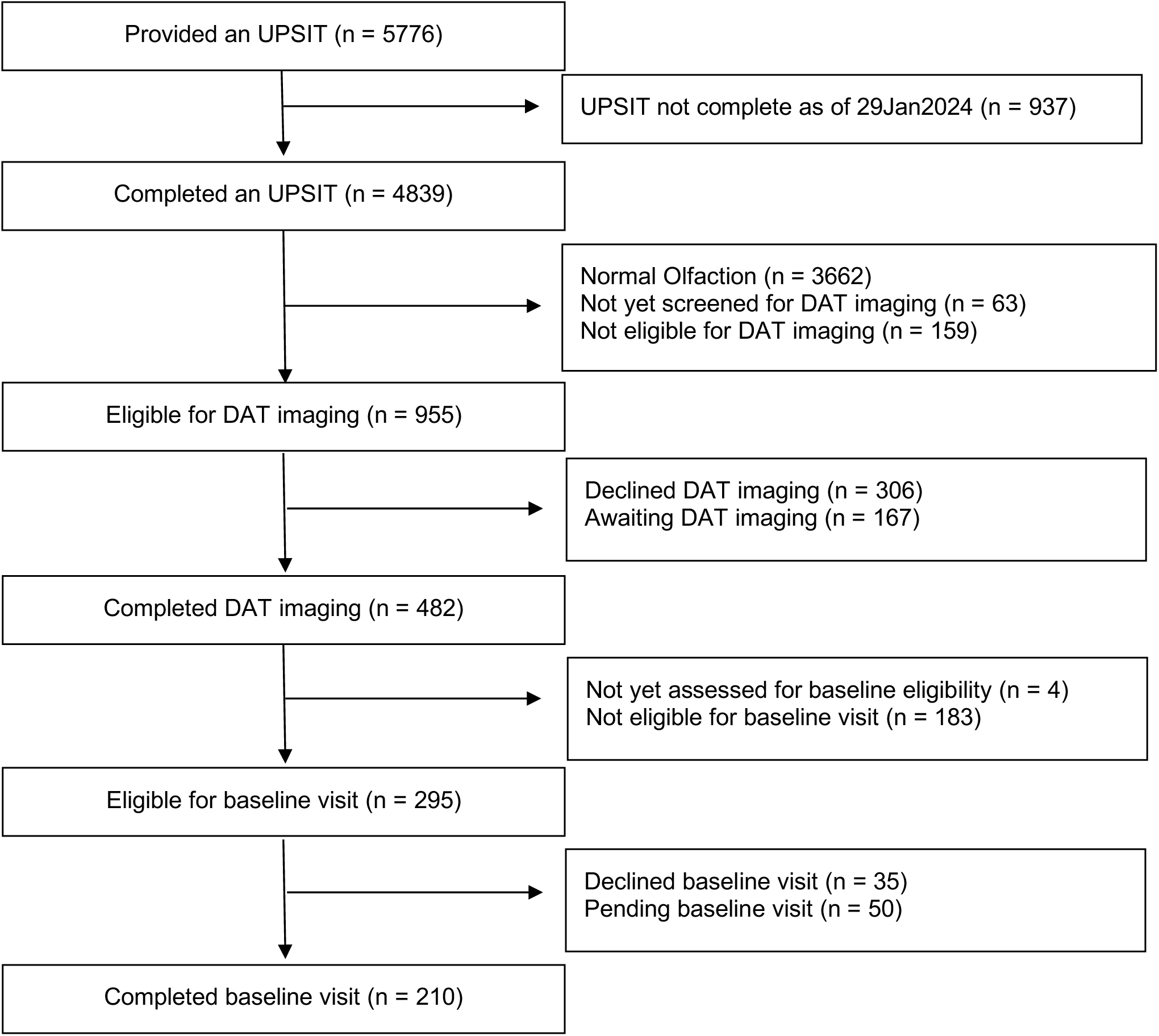
Flowchart of all participants recruited through PPMI Online.

**eFigure 2.**
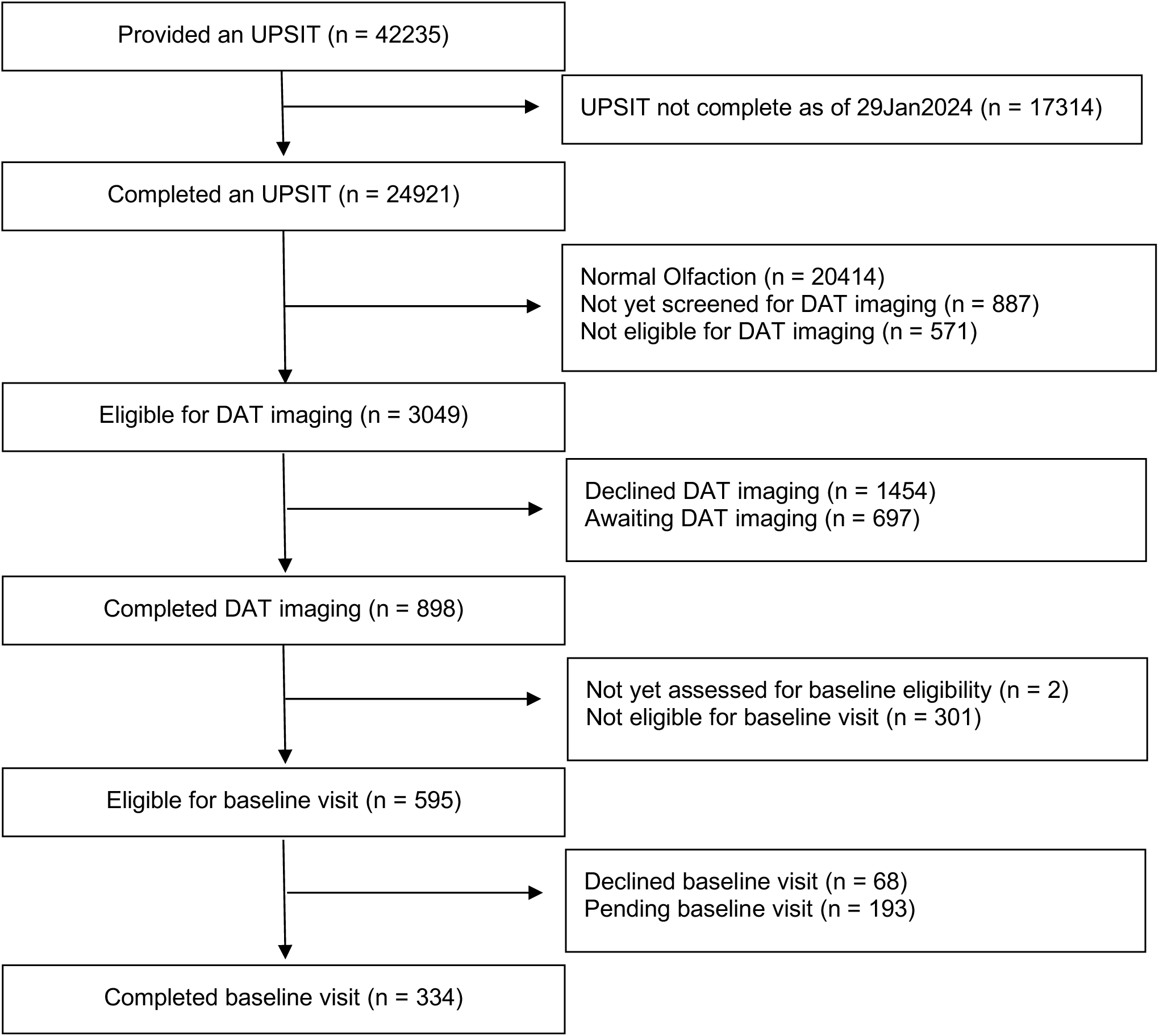
Flowchart of all participants recruited through ST Direct.

**eFigure 3.**
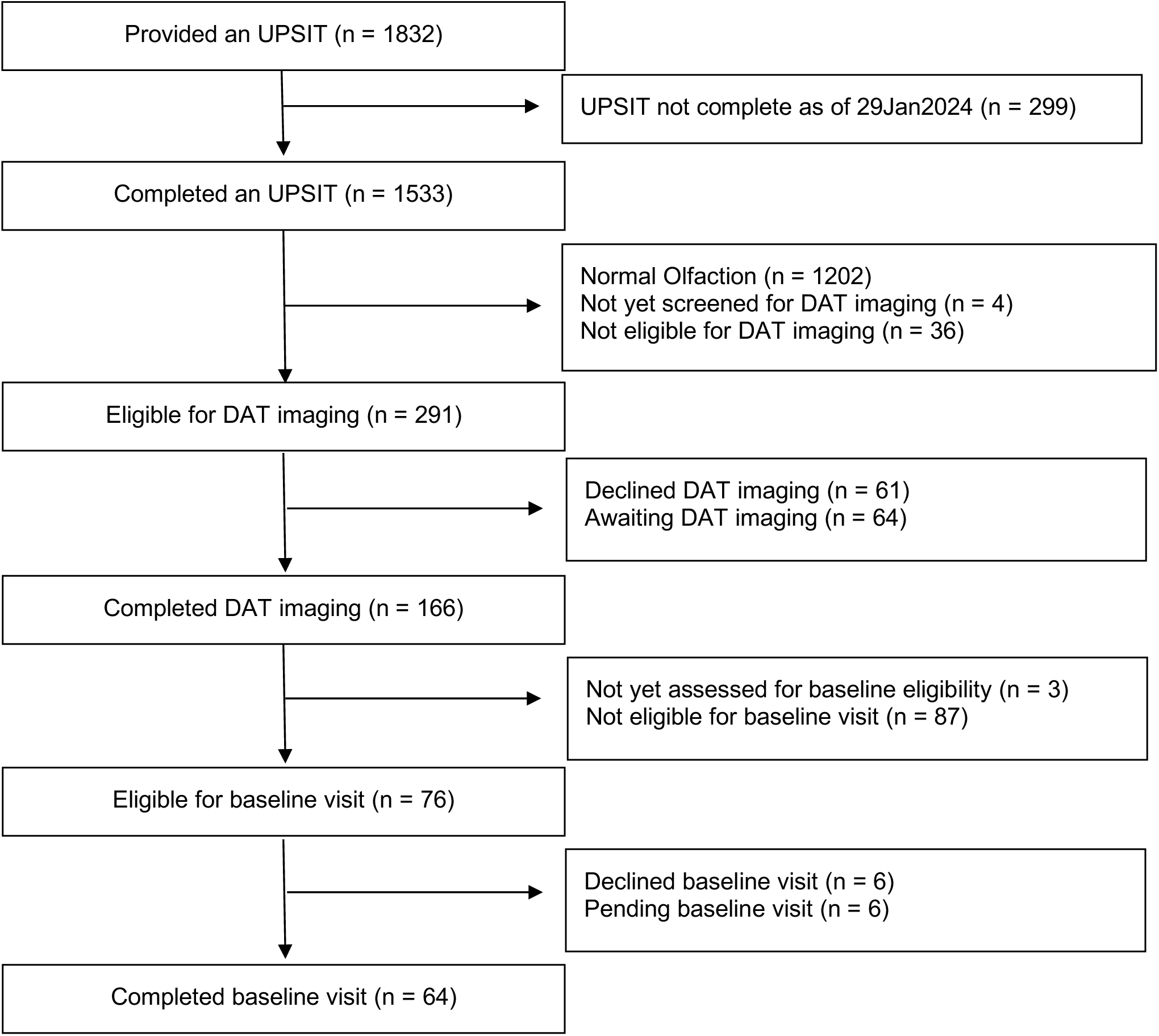
Flowchart of all participants recruited through MJFF Ads.

**eFigure 4:**
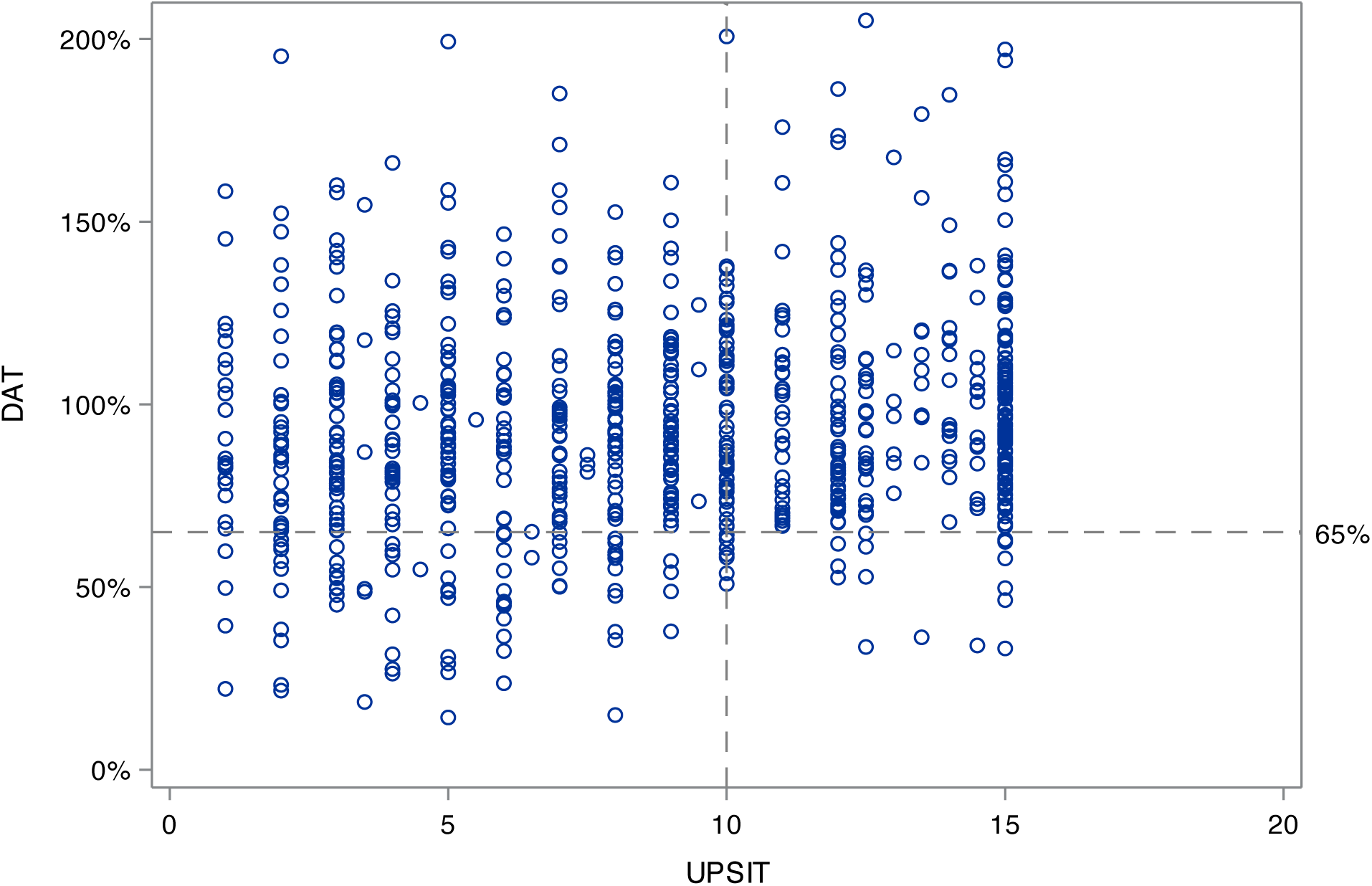
Distribution of UPSIT results and DAT imaging results among participants who completed a smell test prior to January 6^th^, 2023, and were eligible for DAT imaging based on an UPSIT of 15^th^ %ile or less. DAT imaging results shown represent the lowest putamen specific binding ratio divided by the specific binding ratio expected for age and sex.

**eTable 1.** Characteristics of participants who completed UPSIT compared to those that did not complete UPSIT but consented for staged screening.

|  | Participants who completed an UPSIT<br>(N = 31293) | Participants who were provided an UPSIT but did not complete<br>(N = 18550) |
| --- | --- | --- |
| **Age when UPSIT sent (years), Mean (SD)** | 68.2 (6.0) | 67.7 (6.2) |
| Median (Min, Max) | 67.3 (23.6, 95.9) | 66.6 (21.2, 99.8) |
| Missing | 50 | 307 |
| **Sex, n (%)** |  |  |
| Female | 22131 (71%) | 13362 (72%) |
| Male | 9162 (29%) | 5188 (28%) |

**eTable 2:** Characteristics of participants who completed DAT imaging compared to those that declined or were not eligible for DAT imaging, among participants that completed an UPSIT.

|  | Participants that completed DaTScan<br>(N = 1546) | Participants that declined DaTScan<br>(N = 1821) | Participants not eligible for DaTScan<br>(N = 26998) |
| --- | --- | --- | --- |
| **Age at UPSIT (years), Mean (SD)** | 67.7 (5.5) | 69.3 (6.9) | 68.2 (5.9) |
| Median (Min, Max) | 66.9 (59.3, 87.2) | 68.2 (33.9, 95.8) | 67.4 (23.6, 95.9) |
| **Sex, n (%)** |  |  |  |
| Female | 1001 (65%) | 1377 (76%) | 19126 (71%) |
| Male | 545 (35%) | 444 (24%) | 7872 (29%) |
| **UPSIT Percentile, Mean (SD)** | 6.6 (4.0) | 6.0 (3.6) | 33.0 (16.3) |
| Median (Min, Max) | 6.0 (1.0, 15.0) | 5.5 (1.0, 35.0) | 32.0 (1.0, 93.5) |
| **UPSIT Percentile Categories, n (%)** |  |  |  |
| < 10th Percentile | 1221 (79%) | 1608 (88%) | 991 (4%) |
| ≥ 10th and ≤ 15th Percentile | 325 (21%) | 209 (11%) | 3020 (11%) |
| > 15th Percentile | 0 | 4 (<1%) | 22987 (85%) |

**eTable 3.** Responses to select questions given to participants recruited through PPMI Online and ST Direct prior to smell testing. Participants may have endorsed more than one feature.

|  | **PPMI Online**<br>(N = 4839) | **ST Direct**<br>(N = 24921) |
| --- | --- | --- |
| **Diagnosis of RBD, n (%)** | 229 (5%) | 726 (3%) |
| Incomplete, prefer not to answer, or not sure | 202 | 1414 |
| **Self-reported DEB without an RBD diagnosis, n (%)** | 752 (16%) | 2782 (12%) |
| Incomplete, prefer not to answer, or not sure | 191 | 1396 |
| **Self-reported smell loss, n (%)** | 643 (14%) | 4240 (21%) |
| Incomplete, prefer not to answer, or not sure | 334 | 4367 |
| **Self-reported family history, n (%)** | 2264 (49%) | 4759 (20%) |
| Incomplete, prefer not to answer, or not sure | 214 | 1433 |

## PPMI STUDY TEAMS/CORES/COLLABORATORS FOR PUBLICATIONS

### Executive Steering Committee

Kenneth Marek, MD^1^ (Principal Investigator); Caroline Tanner, MD, PhD^9^; Tanya Simuni, MD^3^; Andrew Siderowf, MD, MSCE^12^; Douglas Galasko, MD^27^; Lana Chahine, MD^41^; Christopher Coffey, PhD^4^; Kalpana Merchant, PhD^61^; Kathleen Poston, MD^40^; Roseanne Dobkin, PhD^43^; Tatiana Foroud, PhD^15^; Brit Mollenhauer, MD^8^; Dan Weintraub, MD^12^; Ethan Brown, MD^9^; Karl Kieburtz, MD, MPH^23^; Mark Frasier, PhD^6^; Todd Sherer, PhD^6^; Sohini Chowdhury, MA^6^; Roy Alcalay, MD^36^ and Aleksandar Videnovic, MD^47^

### Steering Committee

Duygu Tosun-Turgut, PhD^9^; Werner Poewe, MD^7^; Susan Bressman, MD^14^; Jan Hammer^15^; Raymond James, RN^22^; Ekemini Riley, PhD^42^; John Seibyl, MD^1^; Leslie Shaw, PhD^12^; David Standaert, MD, PhD^18^; Sneha Mantri, MD, MS^62^; Nabila Dahodwala, MD^12^; Michael Schwarzschild^47^; Connie Marras^45^; Hubert Fernandez, MD^25^; Ira Shoulson, MD^23^; Helen Rowbotham^2^; Paola Casalin^11^ and Claudia Trenkwalder, MD^8^

### Michael J. Fox Foundation (Sponsor)

Todd Sherer, PhD; Sohini Chowdhury, MA; Mark Frasier, PhD; Jamie Eberling, PhD; Katie Kopil, PhD; Alyssa O’Grady; Maggie McGuire Kuhl; Leslie Kirsch, EdD and Tawny Willson, MBS

### Study Cores, Committees and Related Studies: *(Include as applicable to the paper)*

Project Management Core: Emily Flagg, BA^1^

Site Management Core: Tanya Simuni, MD^3^; Bridget McMahon, BS^1^ Strategy and Technical Operations: Craig Stanley, PhD^1^; Kim Fabrizio, BA^1^ Data Management Core: Dixie Ecklund, MBA, MSN^4^; Trevis Huff, BSE^4^

Screening Core: Tatiana Foroud, PhD^15^; Laura Heathers, BA^15^; Christopher Hobbick, BSCE^15^; Gena Antonopoulos, BSN^15^

Imaging Core: John Seibyl, MD^1^; Kathleen Poston, MD^40^

Statistics Core: Christopher Coffey, PhD^4^; Chelsea Caspell-Garcia, MS^4^; Michael Brumm, MS^4^

Bioinformatics Core: Arthur Toga, PhD^10^; Karen Crawford, MLIS^10^

Biorepository Core: Tatiana Foroud, PhD^15^; Jan Hamer, BS^15^

Biologics Review Committee: Brit Mollenhauer^8^; Doug Galasko^27^; Kalpana Merchant^61^

Genetics Core: Andrew Singleton, PhD^13^

Pathology Core: Tatiana Foroud, PhD^15^; Thomas Montine, MD, PhD^40^

Found: Caroline Tanner, MD PhD^9^

PPMI Online: Carlie Tanner, MD PhD^9^; Ethan Brown, MD^9^; Lana Chahine, MD^41^; Roseann Dobkin, PhD^43^; Monica Korell, MPH^9^

Site Investigators:

Charles Adler, PhD^51^; Roy Alcalay, MD^36^; Amy Amara, PhD^52^; Paolo Barone, PhD^30^; Bastiaan Bloem, PhD^60^ Susan Bressman, MD^14^; Kathrin Brockmann, MD^26^; Norbert Brüggemann, MD^59^; Lana Chahine, MD^41^; Kelvin Chou, MD^44^; Nabila Dahodwala, MD^12^; Alberto Espay, MD^32^; Stewart Factor, DO^16^; Hubert Fernandez, MD^25^; Michelle Fullard, MD^52^; Douglas Galasko, MD^27^; Robert Hauser, MD^19^; Penelope Hogarth, MD^17^; Shu-Ching Hu, PhD^21^; Michele Hu, PhD^58^; Stuart Isaacson, MD^31^; Christine Klein, MD^59^; Rejko Krueger, MD^2^; Mark Lew, MD^49^; Zoltan Mari, MD^56^; Connie Marras, PhD^45^; Maria Jose Martí, PhD^34^; Nikolaus McFarland, PhD^54^; Tiago Mestre, PhD^46^; Brit Mollenhauer, MD^8^; Emile Moukheiber, MD^28^; Alastair Noyce, PhD^63^ Wolfgang Oertel, PhD^64^; Njideka Okubadejo, MD^65^; Sarah O’Shea, MD^39^; Rajesh Pahwa, MD^48^; Nicola Pavese, PhD^57^; Werner Poewe, MD^7^; Ron Postuma, MD^55^; Giulietta Riboldi, MD^53^; Lauren Ruffrage, MS^18^; Javier Ruiz Martinez, PhD^35^; David Russell, PhD^1^; Marie H Saint-Hilaire, MD^22^; Neil Santos, BS^51^; Wesley Schlett^47^; Ruth Schneider, MD^23^; Holly Shill, MD^50^; David Shprecher, DO^24^; Tanya Simuni, MD^3^; David Standaert, PhD^18^; Leonidas Stefanis, PhD^38^; Yen Tai, PhD^29^; Caroline Tanner, PhD^9^; Arjun Tarakad, MD^20^; Eduardo Tolosa PhD^34^ and Aleksandar Videnovic, MD^47^

Coordinators:

Susan Ainscough, BA^30^; Courtney Blair, MA^18^; Erica Botting^19^; Isabella Chung, BS^56^; Kelly Clark^24^; Ioana Croitoru^35^; Kelly DeLano, MS^32^; Iris Egner, PhD^7^; Fahrial Esha, BS^53^; May Eshel, MSc^36^; Frank Ferrari, BS^44^; Victoria Kate Foster^57^; Alicia Garrido, MD^34^; Madita Grümmer^59^; Bethzaida Herrera^50^; Ella Hilt^26^; Chloe Huntzinger, BA^52^; Raymond James, BS^22^; Farah Kausar, PhD^9^; Christos Koros, MD, PhD^38^; Yara Krasowski, MSc^60^; Dustin Le, BS^17^; Ying Liu, MD^52^; Taina M. Marques, PhD^2^; Helen Mejia Santana, MA^39^; Sherri Mosovsky, MPH^41^; Jennifer Mule, BS^25^; Philip Ng, BS^45^; Lauren O’Brien^48^; Abiola Ogunleye, PGDip^29^; Oluwadamilola Ojo, MD^65^; Obi Onyinanya, BS^28^; Lisbeth Pennente, BA^31^; Romina Perrotti^55^; Michael Pileggi, MS^55^; Ashwini Ramachandran, MSc^12^; Deborah Raymond, MS^14^; Jamil Razzaque, MS^58^; Shawna Reddie, BA^46^; Kori Ribb, BSN,^28^; Kyle Rizer, BA^54^; Janelle Rodriguez, BS^27^; Stephanie Roman, HS^1^; Clarissa Sanchez, MPH^20^; Cristina Simonet, PhD^29^; Anisha Singh, BS^23^; Elisabeth Sittig, RN^64^; Barbara Sommerfeld MSN^16^; Angela Stovall, BS^44^; Bobbie Stubbeman, BS^32^; Alejandra Valenzuela, BS^49^; Catherine Wandell, BS^21^; Diana Willeke^8^; Karen Williams, BA^3^ and Dilinuer Wubuli, MB^45^

#### Partners Scientific Advisory Board (Acknowledgement)

**Funding: PPMI – a public-private partnership – is funded by the Michael J. Fox Foundation for Parkinson’s Research and funding partners, including 4D Pharma, Abbvie, AcureX, Allergan, Amathus Therapeutics, Aligning Science Across Parkinson’s, AskBio, Avid Radiopharmaceuticals, BIAL, BioArctic, Biogen, Biohaven, BioLegend, BlueRock Therapeutics, Bristol-Myers Squibb, Calico Labs, Capsida Biotherapeutics, Celgene, Cerevel Therapeutics, Coave Therapeutics, DaCapo Brainscience, Denali, Edmond J. Safra Foundation, Eli Lilly, Gain Therapeutics, GE HealthCare, Genentech, GSK, Golub Capital, Handl Therapeutics, Insitro, Jazz Pharmaceuticals, Johnson & Johnson Innovative Medicine, Lundbeck, Merck, Meso Scale Discovery, Mission Therapeutics, Neurocrine Biosciences, Neuron23, Neuropore, Pfizer, Piramal, Prevail Therapeutics, Roche, Sanofi, Servier, Sun Pharma Advanced Research Company, Takeda, Teva, UCB, Vanqua Bio, Verily, Voyager Therapeutics, the Weston Family Foundation and Yumanity Therapeutics.**

1. Institute for Neurodegenerative Disorders, New Haven, CT
2. University of Luxembourg, Luxembourg
3. Northwestern University, Chicago, IL
4. University of Iowa, Iowa City, IA
5. VectivBio AG
6. The Michael J. Fox Foundation for Parkinson’s Research, New York, NY
7. Innsbruck Medical University, Innsbruck, Austria
8. Paracelsus-Elena Klinik, Kassel, Germany
9. University of California, San Francisco, CA
10. Laboratory of Neuroimaging (LONI), University of Southern California
11. BioRep, Milan, Italy
12. University of Pennsylvania, Philadelphia, PA
13. National Institute on Aging, NIH, Bethesda, MD
14. Mount Sinai Beth Israel, New York, NY
15. Indiana University, Indianapolis, IN
16. Emory University of Medicine, Atlanta, GA
17. Oregon Health and Science University, Portland, OR
18. University of Alabama at Birmingham, Birmingham, AL
19. University of South Florida, Tampa, FL
20. Baylor College of Medicine, Houston, TX
21. University of Washington, Seattle, WA
22. Boston University, Boston, MA
23. University of Rochester, Rochester, NY
24. Banner Research Institute, Sun City, AZ
25. Cleveland Clinic, Cleveland, OH
26. University of Tübingen, Tübingen, Germany
27. University of California, San Diego, CA
28. Johns Hopkins University, Baltimore, MD
29. Imperial College of London, London, UK
30. University of Salerno, Salerno, Italy
31. Parkinson’s Disease and Movement Disorders Center, Boca Raton, FL
32. University of Cincinnati, Cincinnati, OH
33. Hospital Clinic of Barcelona, Barcelona, Spain
34. Hospital Universitario Donostia, San Sebastian, Spain
35. Tel Aviv Sourasky Medical Center, Tel Aviv, Israel
36. St. Olav’s University Hospital, Trondheim, Norway
37. National and Kapodistrian University of Athens, Athens, Greece
38. Columbia University Irving Medical Center, New York, NY
39. Stanford University, Stanford, CA
40. University of Pittsburgh, Pittsburgh, PA
41. Center for Strategy Philanthropy at Milken Institute, Washington D.C.
42. 12, New Brunswick, NJ
43. University of Michigan, Ann Arbor, MI
44. Toronto Western Hospital, Toronto, Canada
45. The Ottawa Hospital, Ottawa, Canada
46. Massachusetts General Hospital, Boston, MA
47. University of Kansas Medical Center, Kansas City, KS
48. University of Southern California, Los Angeles, CA
49. Barrow Neurological Institute, Phoenix, AZ
50. Mayo Clinic Arizona, Scottsdale, AZ
51. University of Colorado, Aurora, CO
52. NYU Langone Medical Center, New York, NY
53. University of Florida, Gainesville, FL
54. Montreal Neurological Institute and Hospital/McGill, Montreal, QC, Canada
55. Cleveland Clinic-Las Vegas Lou Ruvo Center for Brain Health, Las Vegas, NV
56. Clinical Ageing Research Unit, Newcastle, UK
57. John Radcliffe Hospital Oxford and Oxford University, Oxford, UK
58. Universität Lübeck, Luebeck, Germany
59. Radboud University, Nijmegen, Netherlands
60. TransThera Consulting
61. Duke University, Durham, NC
62. Wolfson Institute of Population Health, Queen Mary University of London, UK
63. Philipps-University Marburg, Germany
64. University of Lagos, Nigeria

